# Use of Interviewer-Administered Telephone Surveys during Infectious Disease Outbreaks, Epidemics, and Pandemics: A Scoping Review

**DOI:** 10.1101/2022.11.01.22281787

**Authors:** Sayaka Arita, Mouhamadou Faly Ba, Zoumana Traoré, Emmanuel Bonnet, Adama Faye, Valery Ridde

## Abstract

**Introduction:** During the COVID-19 crisis, researchers had to collect data remotely. Telephone surveys and interviews can quickly gather data from a distance without heavy expense. Although interviewer-administered telephone surveys (IATS) can accommodate the needs in international public health research, the literature on its use during infectious disease outbreaks is scarce. This scoping review aimed to map characteristics of IATS during infectious disease outbreaks.

**Methods:** IATS conducted principally during infectious disease outbreaks and answered by informants at least 18 years old were searched from PubMed and EBSCO. There was a manual addition of relevant documents identified during an initial search. Global trends were reported using different groupings, and study details were compared between before and during the COVID-19 pandemic.

**Results:** 70 IATS published between 2003 and 2022 were identified. 57.1 % were conducted during the COVID-19 pandemic. During the COVID-19 pandemic, some changes in the use of this data collection modality were observed. The proportion of IATS in LMICs rose from 3.3 % before the COVID-19 pandemic to 32.5 %. The share of qualitative studies grew from 6.7 % to 32.5 %. IATS performed during the COVID-19 pandemic focused on more diverse, specific population groups, such as patients and healthcare professionals. The usage of mobile phones to do IATS studies increased from 3.3 % to 25.0 %.

**Conclusion:** IATS are used globally with high frequency in the Western Pacific Region and high income countries. During the COVID-19 pandemic, IATS was performed in more countries to investigate more diverse target populations. Nonetheless, researchers should consider how to address technical and financial challenges for ITAS to be more inclusive and representative. For better use and more efficient deployment of IATS, methodological details need to be exchanged.

**What is already known on this topic:**

- Telephones have been playing an important role in data collection especially when data needs to be gathered quickly and remotely during infectious disease outbreaks, humanitarian crises, and natural disasters.
- The use of online surveys is increasing globally alongside digitalization and technological development.
- However, the transformation regarding the use of telephone surveys is not well documented.

**What this study adds:**

- We performed a scoping review to grasp characteristics and trends of telephone surveys.
- We found that more telephone surveys have been conducted in low and middle income countries during the COVID-19 pandemic (32.5 %) compared to before COVID-19 (3.3 %).
- We learned that telephone surveys during the pandemic have investigated more specific and diverse population groups than the pre-pandemic period.

**How this study might affect research, practice or policy:**

- The increased usage of cell phones to operate IATS align with the growing mobile phone ownership, thanks to which the global mobilization of this survey mode might be accelerated in the future.
- However, we observed inadequate information on study details, including the number of languages spoken by interviewers as well as technical enhancement or optimization.
- We encourage sharing techniques and knowledge among researchers whereby ITAS could be further improved and contribute to more inclusive public health research.

## INTRODUCTION

Remote data collection is particularly relevant during infectious disease outbreaks when traditional face-to-face modalities are inappropriate. During the COVID-19 crisis, researchers were obliged to collect data remotely given physical distancing, lockdowns, and travel restrictions to contain the virus. Likewise, in the context of natural disasters and conflicts, and for convenience, it is likely that remote data collection methods will continue to be widely used.

Among several tools, telephones are especially useful to quickly gather both qualitative and quantitative data from a distance without heavy expense[1,2]. This modality tends to have higher response rates too[3]. Phone surveys are also effective to study temporal social contexts in which questions are asked and responded[4]. Respondents have more freedom in answering questions during interviews. In addition, this method easily allows interviewers to develop rapport and build trust[5]. and good interviewers can also ask detailed, complex questions which require clarification[6].

Telephone surveys involving interaction between live interviewers and informants (hereinafter interviewer-administered telephone surveys, IATS) can accommodate the specific needs in international public health research. In many cases, high-income countries (HICs) fund and carry out studies in low-middle-income countries (LMICs), which are more vulnerable and exposed to infectious diseases[7]. Informants from resource-scarce nations may feel vulnerable due to the poverty, widespread illiteracy and linguistic barriers[8] given the linguistic diversity in these countries[9].

Despite the usefulness of IATS during infectious disease outbreaks, the literature on this subject is scarce. Therefore, it is unclear in what context and how this remote data collection method is used in the time of infectious disease outbreaks. This scoping review aimed to identify and map characteristics of IATS responded by informants of at least 18 years old during infectious disease outbreaks.

## METHODS

Whereas a systematic review often addresses a precise question, a scoping review aims to investigate the way research is conducted on a specific subject as well as to identify characteristics of studies[10]. The objective of this research is in line with the purpose of a scoping review, and therefore this type of review was considered appropriate.

### Reflexivity statement

This scoping review intended to produce a global picture illustrating how IATS have been used. Although the first author principally formulated the protocol and performed data searches to conduct this scoping review efficiently, the other authors contributed to screening and selection of documents as well as analysis. By working remotely, the authors from HICs and LMICs were given opportunities to give feedback and exchange insights. There were 1 female and 1 male early career researchers, who took the lead in the scoping review. Given the diversity of the authors’ backgrounds, this scoping review, a product of an international collaboration, can provide insightful ideas for future research and particularly methods, which needs sufficient attention and consideration in global health.

### Protocol and registration

The final version of the protocol[11] was made available on 17 June 2022 online (www.protocols.io/).

### Eligibility criteria

Following the Joanna Briggs Institute Manual for Evidence Synthesis for scoping reviews, the PCC (Population/Participants, Concept, Context) framework was used to identify eligibility criteria. This review included telephone surveys distributed to and responded by adults, anyone at least 18 years old (population/participants). Because this research aims to capture global characteristics and trends of IATS rather than specific socio-demographic factors, sex or gender information was not extracted for all the included studies. Telephone surveys, including both landline and mobile phones, were included. This review was limited to studies which relied on the single method of interviewer-administered telephone surveys (concept). This scoping review included the telephone surveys whose data was collected during infectious disease outbreaks, epidemics, and pandemics (context).

### Information sources and search

The first author (SA) performed database searches in PubMed and EBSCO on 5 April 2022 without restrictions on language or publication date. SA conducted an initial limited search of MEDLINE and found the text words in relevant articles as well as Medical subject Headings (MeSH) terms. Using these words, SA drafted and refined the search strategy in accordance with feedback from a school librarian and other reviewers. To allow the searches to be widest possible, tems like “infectious disease” were used instead of specific illnesses. The following combination was used for the both database searches: (((‘telephone*’OR ‘cellular phone*’ OR ‘phone*’ OR ‘cell phone*’ OR ‘mobile phone*’ OR ‘mobile telephone*’) AND (‘survey*’ OR ‘interview*’ OR ‘cross-sectionalsurvey*’ OR ‘longitudinal survey*’)) OR (‘interviewer-administeredsurvey*’ OR ‘intervieweradministered survey*’ OR ‘computer-assisted telephone interviewing’ OR ‘computer assisted telephone interviewing’)) AND ((‘outbreak*’ OR ‘epidemic*’ OR ‘pandemic*’) AND (‘infectiousdisease*’OR ‘communicablechronicdisease*’ OR ‘communicable infectious disease*’ OR ‘infectious illness’ OR ‘infectious virus*’)).

Additional documents consulted during an initial search prior to the protocol development were also added as potentially relevant studies. Both a protocol and a search strategy were developed by the corresponding author and agreed by the other reviewers.

### Selection of sources of evidence

Two reviewers (SA and MFB) independently screened the potentially relevant documents using a web-based platform, Covidence (www.covidence.org/). Duplicates were removed automatically. Due to the reviewers’ language proficiency, studies were excluded if not written in English, French, Spanish, or Japanese. Any disagreements were resolved by discussions.

### Data charting process and items

Using Microsoft Excel 16.61.1, the first author drafted a data extraction form. This form was reviewed by the other authors and continuously updated as needed. Data was extracted by the corresponding author, which was later reviewed and verified by the other reviewers. Information extracted from each study includes general information and study details, and a full list of the extracted items can be found in Table 1. The included studies were sorted in alphabetical order by the corresponding author’s last name.

**Table 1.** List of Items Extracted from Included Studies

| **(A) Basic Information** |  |
| --- | --- |
| Authors | Corresponding author's name followed by et al. |
| Year | Year of publication. |
| Title | Title of the publication. |
| Journal | Journal in which the study is published. |
| Origin | Country/countries in which the data is collected. |
| Purpose | Study objective(s). |
| Sample Size | Number of observations included in the final analysis. |
| Design | Categorized based on Mixed Methods Appraisal Tool (MMAT).(12)<br>1. Qualitative<br>2. Quantitative randomized controlled trials<br>3. Quantitative non-randomized<br>4. Quantitative descriptive<br>5. Mixed Methods |
| **(B) Study Details** |  |
| Scale | Categorized based on the scale of data collection.<br>1. International if the study uses data gathered in multiple countries;<br>2. National if the study sample was deemed to be nationally representative and when there is no geographical limitation to draw samples;<br>3. Regional if the data was collected from multiple states or prefectures;<br>4. Local if the data came from a single state or prefecture. |
| Population | Data related to the target population was extracted and categorized into 5 groups.<br>1. Adult individuals if the study concerns the general population, or anyone at least 18 years old;<br>2. Healthcare professionals (HCPs) if the study targets healthcare providers, medical practitioners, and other hospital staff;<br>3. Patients if the IATS focuses on patients of any disease;<br>4. Households if the study was designed to collect household-level data, and the sample size for this group is the number of households included in the final analysis;<br>5. Other if the target population does not fall into any of the groups described above, and when the target population is a combination of the above-mentioned groups. |
| Infectious Disease | Infectious disease(s) present during data collection. |
| Phone type | The type of telephone used in each study was assessed and grouped into 4 categories.<br>1. Landline if the IATS uses exclusively or largely home landlines, fixed lines, and residential telephones; |
|  | 2. Mobile if the data was collected solely or predominantly by mobile phones, including smartphones;<br>3. 50-50 if both landlines and mobiles phones were used at the ratio 1 to 1;<br>4. Not specified if the telephone type was not specified, or when the share of each phone type was not mentioned. |

### Critical appraisal of individual sources of evidence

We appraised neither methodological quality nor risk of bias of the included studies. This is consistent with guidance for conducting a scoping review[13]. However, the MMAT version 2018[12] was employed to determine the study design. The quality or the ratings were not presented.

### Synthesis of results

The reporting of this scoping review follows the PRISMA-ScR guidelines[13]. Considering that the prevalence of infectious diseases varies in different countries and regions, the included IATS were grouped geographically (WHO region) for descriptive analyses. Furthermore, given that LMICs are more likely to be exposed to infectious diseases and study sites than HICs, a financial grouping (World Bank income classification) was also used to detect trends in the use of telephone surveys. RStudio 4.2.1 was used for the descriptive analyses.

## RESULTS

### Selection of sources of evidence

A total of 526 records were retrieved via database searches, and 4 documents were added as supplemental studies. After removing duplicates, 420 potential studies were screened by title and abstract. Of these, 146 were screened by full text, and 70 were included for the analysis. Of these, 66 studies were identified via database searches, and 4 were manually added. All studies are written in English. The screening process can be found in Figure 1.

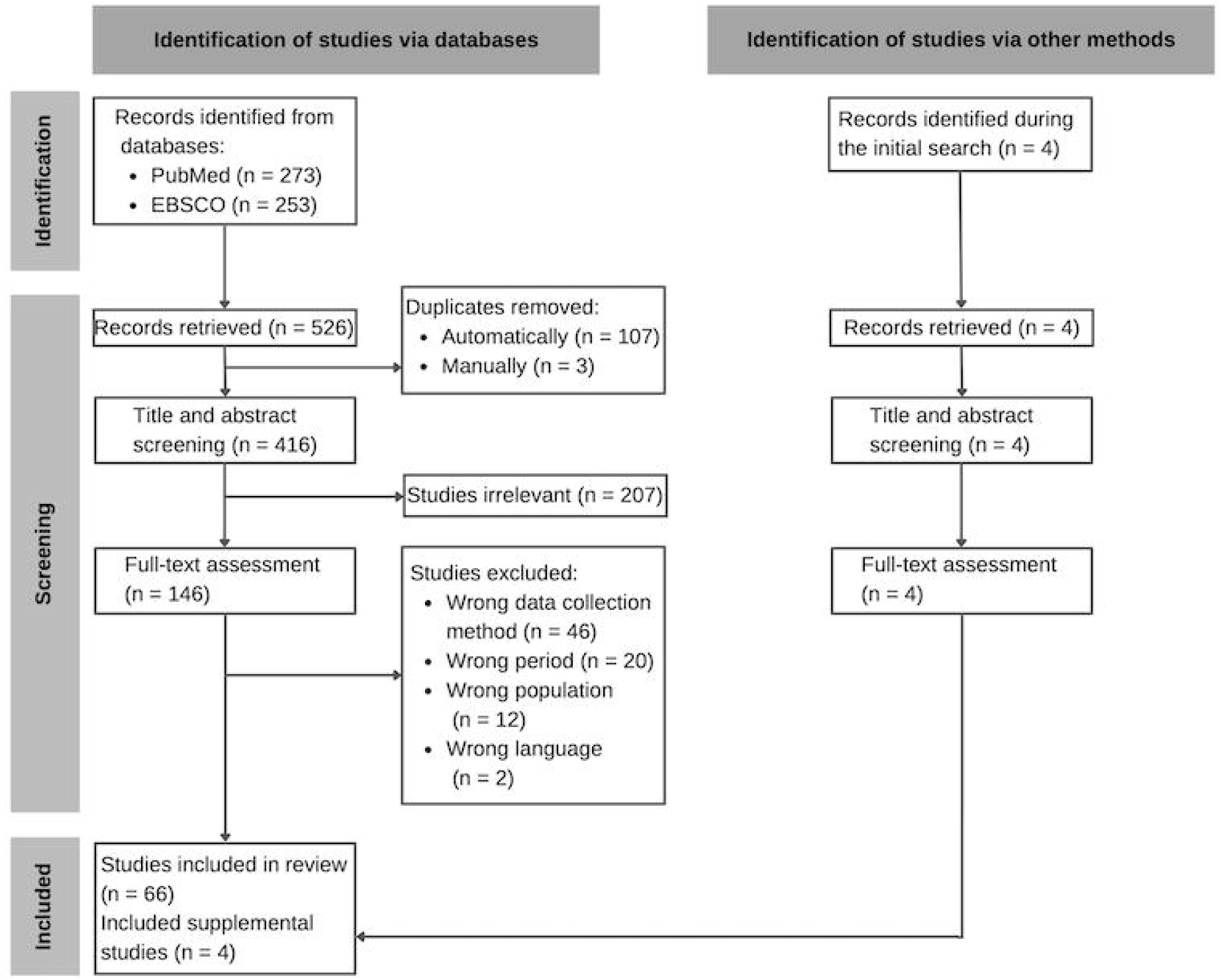

### Characteristics of sources of evidence

This scoping review included 70 IATS published between 2003 and 2022 from all over the world. The included studies were performed during the following infectious disease outbreaks: Chikungunya, COVID-19, Dengue, Ebola, H1N1, H5N1, H7H9, SARS, Seasonal flu, and Zika. There was an upsurge in the number of IATS published during the COVID-19 pandemic.

### Results of individual sources of evidence

A full list of all the included studies in this scoping review can be found in Supplemental Materials 1 and 2.

### Synthesis of results

The 70 studies were found in all the 6 WHO regions as seen in Figure 2. 33 studies (47.1%) were found in the Western Pacific Region (WPRO), 13 (18.6%) in the European Region (EURO), 10 (14.3%) in the Region of the Americas (AMRO), 10 (14.3%) in the African Region (AFRO), 2 (2.9%) in the Eastern Mediterranean Region (EMRO), and 2 (2.9%) in the South-East Asia Region (SEARO).

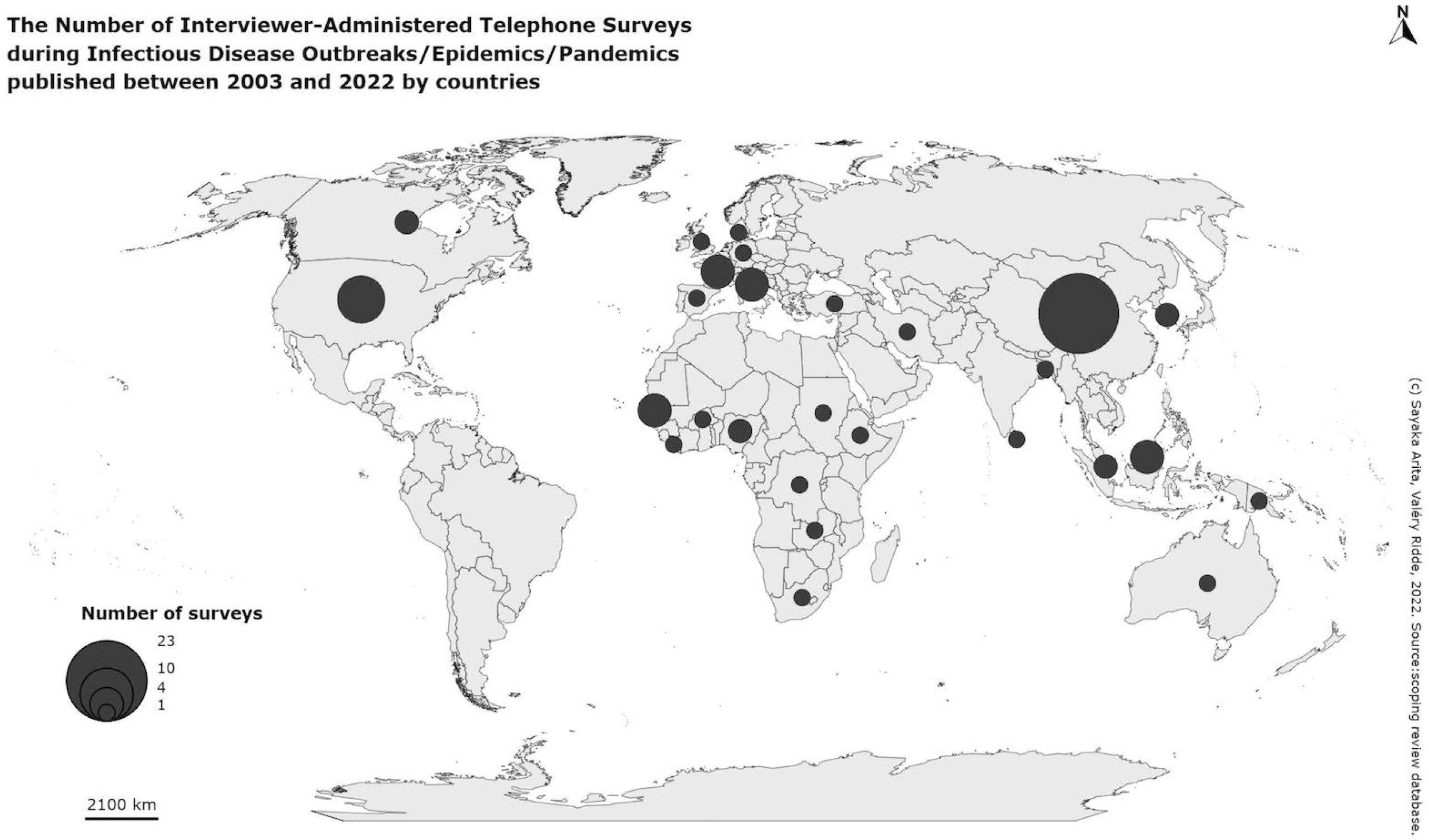

Among the 70 studies, 27 (38.6%) were found in high income countries, 29 (41.4%) in upper-middle income countries, 10 (14.3%) in lower-middle income countries, and 3 (4.3%) in low income countries. In addition, there was 1 international study (1.4%) performed in low and lower-middle income countries (Burkina Faso, Ethiopia, and Nigeria). These income groups can be further merged and simplified. If high and upper-middle income countries are combined as high income countries (HICs), and low and lower-middle income countries are put together as LMICs, 56 (80.0 %) studies were found in HICs and 14 (20.0 %) in LMICs.

In the 70 studies, the smallest sample size was 11(14), and the largest was 31332(15). There were 35 quantitative descriptive studies (50.0 %), 16 quantitative non-randomized studies (22.9 %), 15 qualitative studies (21.4 %), 3 mixed methods studies (4.3 %), and 1 quantitative randomized controlled trial (1.4 %). In terms of the scale, 35 studies were at the local level (50.0 %), 22 regional (31.4 %), 12 national (17.1 %), and 1 international (1.4 %).

Among all the included studies, adult individuals were the most surveyed population with 34 studies (48.6 %). There were 10 IATS targeting patients (14.3 %), 7 HCPs (10.0 %), 7 households (10.0 %), and 12 other groups (17.1 %). Although 35 studies did not specify the phone type (50.0 %), 21 studies relied on landlines (30.0 %). 11 studies used mobile phones (15.7 %), and 3 studies used fixed and cell phones at the ratio of 1 to 1 (4.3 %).

Of 70 IATS, 40 (57.1 %) were carried out during the COVID-19 pandemic, 1 of which was conducted in the Democratic Republic of the Congo where an outbreak of Ebola was also declared[16]. There were 30 studies which took place before the COVID-19 pandemic, and 17 were realized during the H1N1 pandemic, 1 of which compared social-cognitive factors on personal hygiene practices between the H1N1 and H5N1 outbreaks in Hong Kong[17]. All the infectious diseases during which the included IATS were carried out are listed in Table 2.

**Table 2.** Number of IATS by Infectious Disease; n (%)

|  |  |  |
| --- | --- | --- |
| During COVID-19 Pandemic<br>(n = 40) | COVID-19 | 39 (55.7 %) |
|  | COVID-19, Ebola | 1 (1.4 %) |
| Before COVID-19 Pandemic<br>(n = 30) | Chikungunya, Dengue, Zika | 1 (1.4 %) |
|  | Dengue | 1 (1.4 %) |
|  | Ebola | 2 (2.9 %) |
|  | H1N1 | 16 (22.9 %) |
|  | H1N1, H5N1 | 1 (1.4 %) |
|  | H7N9 | 1 (1.4 %) |
|  | SARS | 3 (4.3 %) |
|  | Seasonal Flu | 2 (2.9 %) |
|  | Zika | 3 (4.3 %) |

The use of IATS greatly increased during the COVID-19 pandemic, and some characteristics changed since the pre-pandemic period as seen in Table 3. For example, there were no IATS conducted in EMRO and SEARO before the COVID-19 pandemic. However, during the pandemic, this remote data collection method was mobilized in all WHO regions. WPRO remains the region where this method has been most frequently used. Furthermore, the use of this data collection mode in LMICs rose from 3.3 % before the COVID-19 pandemic to 32.5 % during the pandemic.

**Table 3.** Characteristics of IATS Before and During COVID-19 Pandemic

|  | Before COVID-19 Pandemic<br>(n = 30) | During COVID-19 Pandemic<br>(n = 40) |
| --- | --- | --- |
| WHO Region |  |  |
| African Region (AFRO) | 1 (3.3 %) | 9 (22.5 %) |
| Region of the Americas (AMRO) | 5 (16.7 %) | 5 (12.5 %) |
| Eastern Mediterranean Region (EMRO) | 0 (0.0 %) | 2 (5.0 %) |
| European Region (EURO) | 4 (13.3 %) | 9 (22.5 %) |
| South-East Asia Region (SEARO) | 0 (0.0 %) | 2 (5.0 %) |
| Western Pacific Region (WPRO) | 20 (66.7 %) | 13 (32.5 %) |
| HICs and LMICs |  |  |
| HICs | 29 (96.7 %) | 27 (67.5 %) |
| LMICs | 1 (3.3 %) | 13 (32.5 %) |
| Sample Size |  |  |
| Minimum | 28 | 11 |
| Maximum | 12965 | 31332 |
| Median | 1050 | 412.5 |
| Mean | 2231.3 | 2518 |
| Study Design |  |  |
| Qualitative | 2 (6.7 %) | 13 (32.5 %) |
| Quantitative Randomized Controlled Trials | 0 (0.0 %) | 1 (2.5 %) |
| Quantitative Non-Randomized | 8 (26.7 %) | 8 (20.0 %) |
| Quantitative Descriptive | 20 (66.7 %) | 15 (37.5 %) |
| Mixed Methods | 0 (0.0 %) | 3 (7.5 %) |
| Scale |  |  |
| International | 0 (0.0 %) | 1 (2.5 %) |
| National | 2 (6.7 %) | 10 (25.0 %) |
| Regional | 11 (36.7 %) | 11 (27.5 %) |
| Local | 17 (56.7 %) | 18 (45.0 %) |
| Population |  |  |
| Adult Individuals | 21 (70.0 %) | 13 (32.5 %) |
| Households | 6 (20.0 %) | 1 (2.5 %) |
| HCPs | 0 (0.0 %) | 7 (17.5 %) |
| Patients | 0 (0.0 %) | 10 (25.0 %) |
| Other | 3 (10.0 %) | 9 (22.5 %) |
| Phone Type |  |  |
| Landline | 20 (66.7 %) | 1 (2.5 %) |
| Mobile | 1 (3.3 %) | 10 (25.0 %) |
| 50-50 | 1 (3.3 %) | 2 (5.0 %) |
| Not Specified | 8 (26.7 %) | 27 (67.5 %) |

There were other aspects that altered during the global outbreak of COVID-19. The range of sample sizes widened. The share of qualitative studies grew from 6.7 % to 32.5 %. Whereas local studies continue to be the dominant study scale both pre- and during COVID-19 pandemic periods (56.7 % and 45.0 % respectively), the share of national studies expanded from 6.7 % to 25.0 %. In terms of the population, before the pandemic, 70.0 % of the IATS targeted adult individuals. However, during the pandemic, IATS focused on more diverse, specific population groups, such as patients (25.0 %) and HCPs (17.5 %), who were not surveyed prior to the pandemic. The share of studies concerning adult individuals decreased to 32.5 % in the time of COVID-19. Lastly, the composition of phone types transformed too. Whilst 66.7 % of the pre-pandemic IATS relied on fixed phones, the share of landline studies shrank to 2.5 % during the COVID-19 outbreak. On the contrary, mobile studies increased from 3.3 % to 25.0 %.

## DISCUSSION

### Summary of evidence

The findings demonstrated that IATS have been carried out around the world with greater frequency in WPRO. Although having been mobilized in restricted regions before the COVID-19 pandemic, this remote data collection method was used more globally amid the pandemic.

This phenomenon corresponds to the fact that COVID-19 was spread and studied internationally whereas other infectious diseases were present more locally and regionally. The expanded usage of cell phones to administer IATS during the COVID-19 outbreak accords with the recent technological advancement particularly in LMICs. Thus, the technological development might have led to the wider use of IATS amid the COVID-19 pandemic to some degree. The same number of IATS performed in AFRO and EURO during the worldwide COVID-19 outbreak appears to hint at the suitability of this remote data collection method during infectious disease outbreaks regardless of the geographical location or income level. It is possible that a further expansion of network coverage and affordability of mobile phones, thanks to which cell phone ownership is increasing in LMICs[18], will accelerate the global mobilization of IATS in the future.

On the other hand, before the COVID-19 outbreak, a lot fewer IATS have been carried out in LMICs even though these countries are more affected and exposed to infectious diseases. In other words, this data collection method was not used in LMICs as commonly as in HICs prior to the COVID-19 pandemic. The less frequent use of IATS prior to the COVID-19 pandemic, despite the higher prevalence of other infectious diseases, might indicate that public health research in LMICs is not sufficient. Or this can be stated that other data collection modalities were preferred and appropriate in LMICs before the COVID-19 outbreak. As discussed earlier, it seems that the rapid growth of mobile phone ownership contributes to spurring the wider use of IATS globally. However, phone ownership is not universal, and technical and financial obstacles continue to exist.

There are different approaches to address technical and financial challenges, such as optimization and technological enhancement. Sharing of techniques and knowledge is encouraged to ameliorate IATS in resource scarce settings, like LMICs. However, the included studies lacked information on techniques unique to telephone surveys, including computer-assisted telephone interviewing (CATI), a Reactive Auto Dialer (RAD), random-digit dialing (RDD), and short message service (SMS). CATI signifies any computer-supported feature of telephone interviews, both hardware and software, which can be a sole part or combined components[19]. RAD allows automatic and optimized calls[20]. RDD is a form of probability sampling by randomly choosing phone numbers[19]. In several included studies, SMS was used to contact potential informants[20–24]. Knowledge sharing seems practical for strategizing and running IATS in a novel, complex circumstance, notably in LMICs where there are individuals with several cell phone subscriptions[9,25].

The new situation regarding phone ownership[18] makes it difficult to accurately estimate the degree of representativeness and the characteristics of (non-)respondents too. As often seen in the included IATS, demographic information is essential when understanding and interpreting the results. Furthermore, socio-economic factors might help more accurately comprehend the survey answers. Nonetheless, it is not always easy to obtain and verify socio-economic variables in LMICs. When relatively recent, reliable data, like censuses, is not accessible as a reference, researchers might need to consider narrowing down the target population, rather than trying to achieve a nationally representative sample of the general public. Having a narrower target population would enable researchers to estimate representativeness of their samples more accurately, thereby the reliability of their data would also be ensured.

Researchers should also pay attention to inclusiveness. As highlighted in some studies, it is important to consider who are included in and excluded from telephone surveys. Whereas some argue that mobile phones can be useful to communicate with harder-to-reach sub-groups[9], others assert that the most marginalized, including those without a stable signal or any form of telephone, are often excluded from research[26–28]. There are several methods to make research more inclusive. For instance, thanks to the low-price of phones, when investigating or attempting to include the most disadvantaged or people without telephone in rural areas, researchers can distribute affordable cell phones to the selected respondents[6] although this solution requires ethical, technical, and financial consideration. Another solution to avoid participants’ financial burdens, especially when targeting the disadvantaged, is using toll-free numbers or offering financial compensation.

Understanding and accommodating local needs is vital to more inclusive research and efficient deployment of IATS too. As many of IATS were at the local level, it is also important to acknowledge the context specific to the study site. To do so, it is crucial to include local staff and experts, who not only reinforce localized knowledge helpful to plan surveys and better comprehend the results but also communicate with informants. Recruitment of multilingual locals and training of interviewers can facilitate building rapport and trust between operators and informants. Among the included studies, the biggest number of languages spoken by interviewers was 6 in 3 studies in Senegal[21,22,28]. Coupled with cultural and linguistic appreciation and proficiency, more personal and direct communication between interviewers and informants in IATS can contribute to minimizing miscommunication and misunderstanding. To answer phone surveys, respondents do not need to be literate or have internet access. IATS also allow probing and clarification. These are strengths of this data collection method when gathering in-depth, qualitative data and studying public health topics in LMICs. However, most of the included studies lacked sufficient information on languages spoken by interviewers. More information on consideration and arrangements for IATS in LMICs needs to be exchanged among researchers.

Lastly, adequate consideration for questions themselves is also the key to good IATS. For instance, characteristics of the question influences item nonresponse since to some extent[29]. It should also be remembered that keeping surveys or interviews short (no longer than about 20-30 minutes)[6,30] can contribute to minimizing informants’ fatigue[2]. This is especially important when using IATS for qualitative research, which tends to involve lengthy interviews. In some of the 15 qualitative studies identified in this scoping review, the average or median length of each session was prone to be long (over 40 minutes)[31–35].

### Limitations

To make this scoping review feasible, studies using multiple or hybrid data collection methods like online questionnaires and meetings in addition to interviewer-administered surveys were excluded although these methods are becoming ubiquitous. Furthermore, only 2 databases were searched, and all the included studies were published in English.

## CONCLUSION

The included studies demonstrated several changes in the use of IATS during the COVID-19 pandemic. For instance, ITAS were concentrated in WPRO, EURO, and HICs prior to the COVID-19 pandemic, but the use of this data collection method increased and spread to more counties, particularly in AFRO and LMICs, during the COVID-19 pandemic. Other interesting changes observed include the more diverse target population as well as the increase of qualitative and national studies. These findings seem to indicate that ITAS is useful during infectious disease outbreaks regardless of the geographical location or income level.

On the other hand, we believe that this data collection modality can be further improved if researchers share more techniques and knowledge by detailing their methods when publishing their studies. We recognized the upsurge of ITAS during the COVID-19 outbreak, notably in LMICs. Together with the complexity and expansion of mobile phone ownership, researchers who intend to do phone surveys and interviews in resource scarce settings need to prepare and strategize their studies carefully. Special consideration for the hard-to-reach groups and the most disadvantaged is necessary too.

For future public health research to be more inclusive and representative, it is important to understand and accommodate local needs, such as linguistic and cultural diversity, by recruiting experts or operators who know the context specific to the study site, for example. Moreover, like other data collection modes, researchers should try to mitigate any financial burden and distress incurred by ITAS. The large number of telephone surveys nowadays can cause pressure on respondents. This pressure might further lead to more informants’ refusal to participate in IATS. Furthermore, questions should be carefully formulated and structured so that higher response rates can be achieved. It will be interesting to see the trend in response rates in future research.

## Data Availability

All data produced in the present study are available upon reasonable request to the authors.

## ACKNOWLEDGEMENTS

This research is a scoping review concerning the existing literature. Therefore, no patients or human subjects were involved in this research, and it was not necessary to obtain any approval by the ethics committee.

I would like to thank Lola Traverson for her insightful comments and suggestions that I received while drafting a protocol. I would also like to recognize the assistance of Flore-Apolline Roy, who created the world map demonstrating where IATS have been conducted.

## REFERENCES

1. Burnard P. The telephone interview as a data collection method. Nurse Educ Today. 1994 Feb 1;14(1):67–72.

2. Hoogeveen J, Pape U. Data Collection in Fragile States: Innovations from Africa and Beyond [Internet]. Washington, DC: World Bank; 2020 [cited 2022 Feb 21]. Available from: https://openknowledge.worldbank.org/handle/10986/32576

3. Jones T, Baxter M, Khanduja V. A quick guide to survey research. Ann R Coll Surg Engl. 2013 Jan;95(1):5–7.

4. Kempf AM, Remington PL. New Challenges for Telephone Survey Research in the Twenty-First Century. Annu Rev Public Health. 2007 Apr 1;28(1):113–26.

5. Hensen B, Mackworth-Young CRS, Simwinga M, Abdelmagid N, Banda J, Mavodza C, et al. Remote data collection for public health research in a COVID-19 era: ethical implications, challenges and opportunities. Health Policy Plan. 2021 Apr 1;36(3):360–8.

6. Croke K, Dabalen A, Demombynes G, Giugale M, Hoogeveen J. Collecting high frequency panel data in Africa using mobile phone interviews. The World Bank; 2012 Jun p. 29. Report No.: 6079.

7. Boutayeb A. The Burden of Communicable and Non-Communicable Diseases in Developing Countries. Handb Dis Burd Qual Life Meas. 2010;531–46.

8. Marshall PA. Module 3: Public Health Research and Practice in International Settings: Special Ethical Concerns. In: Jennings B, Khan J, Mastroianni A, Parker LS, editors. Ethics and Public Health: Model Curriculum. Association of Schools of Public Health; 2003. p. 85–102.

9. Ali J, Labrique AB, Gionfriddo K, Pariyo G, Gibson DG, Pratt B, et al. Ethics Considerations in Global Mobile Phone-Based Surveys of Noncommunicable Diseases: A Conceptual Exploration. J Med Internet Res. 2017 May 5;19(5):e7326.

10. Munn Z, Peters MDJ, Stern C, Tufanaru C, McArthur A, Aromataris E. Systematic review or scoping review? Guidance for authors when choosing between a systematic or scoping review approach. BMC Med Res Methodol. 2018 Nov 19;18(1):143.

11. Arita S, Ba MF, Traoré Z, Bonnet E, Faye A, Ridde V. Use of Interviewer-Administered Telephone Surveys during Infectious Disease Outbreaks, Epidemics, and Pande… [Internet]. protocols.io. 2022 [cited 2022 Sep 5]. Available from: https://www.protocols.io/view/use-of-interviewer-administered-telephone-surveys-cbg6sjze

12. Hong QN, Pluye P, Fàbregues S, Bartlett G, Boardman F, Cargo M, et al. Mixed Methods Appraisal Tool (MMAT), version 2018. 1148552.

13. Tricco AC, Lillie E, Zarin W, O’Brien KK, Colquhoun H, Levac D, et al. PRISMA Extension for Scoping Reviews (PRISMA-ScR): Checklist and Explanation. Ann Intern Med. 2018 Oct 2;169(7):467–73.

14. Abba MA, Badaru UM, Mukhtar NB, Abdullahi A, Mohammed J. Experiences of Healthcare Workers of Hospitalised Individuals with covid-19 in Kano, Nigeria: A Qualitative Study. Afr Focus. 2021 Dec 14;34(2):323–42.

15. Gao H, Du Z, Tsang TK, Xiao J, Shan S, Liao Q, et al. Pandemic fatigue and attenuated impact of avoidance behaviours against COVID-19 transmission in Hong Kong by cross-sectional telephone surveys. BMJ Open. 2021 Dec 1;11(12):e055909.

16. Stoop N, Desbureaux S, Kaota A, Lunanga E, Verpoorten M. Covid-19 vs. Ebola: Impact on households and small businesses in North Kivu, Democratic Republic of Congo. World Dev. 2021 Apr 1;140:105352.

17. Liao Q, Cowling BJ, Lam WWT, Fielding R. The Influence of Social-Cognitive Factors on Personal Hygiene Practices to Protect Against Influenzas: Using Modelling to Compare Avian A/H5N1 and 2009 Pandemic A/H1N1 Influenzas in Hong Kong. Int J Behav Med. 2011;18(2):93–104.

18. Greenleaf AR, Gibson DG, Khattar C, Labrique AB, Pariyo GW. Building the Evidence Base for Remote Data Collection in Low- and Middle-Income Countries: Comparing Reliability and Accuracy Across Survey Modalities. J Med Internet Res. 2017 May 5;19(5):e140.

19. Wolter K, Chowdhury S, Kelly J. Chapter 7 -Design, Conduct, and Analysis of Random-Digit Dialing Surveys. In: Rao CR, editor. Handbook of Statistics [Internet]. Elsevier; 2009 [cited 2022 May 30]. p. 125–54. (Handbook of Statistics; vol. 29). Available from: https://www.sciencedirect.com/science/article/pii/S0169716108000072

20. Diallo AI, Faye A, Tine JAD, Ba MF, Gaye I, Bonnet E, et al. Factors associated with the acceptability of government measures to address COVID-19 in Senegal. Rev DÉpidémiologie Santé Publique. 2022 Jun 1;70(3):109–16.

21. Ridde V, Ba MF, Gaye I, Diallo AI, Bonnet E, Faye A. Participating in a vaccine trial for COVID-19 in Senegal: trust and information. Hum Vaccines Immunother. 2021 Nov 2;17(11):3907–12.

22. Ridde V, Kane B, Gaye I, Ba MF, Diallo A, Bonnet E, et al. Acceptability of government measures against COVID-19 pandemic in Senegal: A mixed methods study. PLOS Glob Public Health. 2022 Apr 25;2(4):e0000041.

23. Kuehne A, Lynch E, Marshall E, Tiffany A, Alley I, Bawo L, et al. Mortality, Morbidity and Health-Seeking Behaviour during the Ebola Epidemic 2014–2015 in Monrovia Results from a Mobile Phone Survey. PLoS Negl Trop Dis. 2016 Aug 23;10(8):e0004899.

24. Smaghi BS, Collins J, Dagina R, Hiawalyer G, Vaccher S, Flint J, et al. Barriers and enablers experienced by health care workers in swabbing for COVID-19 in Papua New Guinea: A multi-methods cross-sectional study. Int J Infect Dis. 2021 Oct;110:S17–24.

25. Le Nestour A, Mbaye S, Moscoviz L. Enquête téléphonique sur la crise du Covid au Sénégal. Cent Glob Dev. 2020 Apr 23;25.

26. Demombynes G, Gubbins P, Romeo A. Challenges and opportunities of mobile phone-based data collection: Evidence from South Sudan. The World Bank; 2013.

27. Gibson DG, Pereira A, Farrenkopf BA, Labrique AB, Pariyo GW, Hyder AA. Mobile Phone Surveys for Collecting Population-Level Estimates in Low- and Middle-Income Countries: A Literature Review. J Med Internet Res. 2017 May 5;19(5):e139.

28. Ba MF, Faye A, Kane B, Diallo AI, Junot A, Gaye I, et al. Factors associated with COVID-19 vaccine hesitancy in Senegal: a mixed study. Hum Vaccines Immunother. 2022 May 11;1–12.

29. Lipps O, Monsch GA. Effects of Question Characteristics on Item Nonresponse in Telephone and Web Survey Modes. Field Methods. 2022 Aug 28;1525822X221115838.

30. Adjiwanou V, Alam N, Alkema L, Asiki G, Bawah A, Béguy D, et al. Measuring excess mortality during the COVID-19 pandemic in low- and lower-middle income countries: the need for mobile phone surveys [Internet]. SocArXiv; 2020 [cited 2022 Feb 21]. Available from: https://osf.io/preprints/socarxiv/4bu3q/

31. Abdelmagid N, Ahmed SAE, Nurelhuda N, Zainalabdeen I, Ahmed A, Fadlallah MA, et al. Acceptability and feasibility of strategies to shield the vulnerable during the COVID-19 outbreak: a qualitative study in six Sudanese communities. BMC Public Health. 2021 Dec;21(1):1153.

32. Lee N, Lee HJ. South Korean Nurses’ Experiences with Patient Care at a COVID-19-Designated Hospital: Growth after the Frontline Battle against an Infectious Disease Pandemic. Int J Environ Res Public Health. 2020 Dec 3;17(23):9015.

33. Mwamba C, Kerkhoff AD, Kagujje M, Lungu P, Muyoyeta M, Sharma A. Diagnosed with TB in the era of COVID-19: patient perspectives in Zambia. Public Health Action. 2020 Dec 21;10(4):141–6.

34. Rubin GJ, Harper S, Williams PD, Öström S, Bredbere S, Amlôt R, et al. How to support staff deploying on overseas humanitarian work: a qualitative analysis of responder views about the 2014/15 West African Ebola outbreak. Eur J Psychotraumatology. 2016 Dec 1;7(1):30933.

35. Kirk Sell T, Ravi SJ, Watson C, Meyer D, Pechta LE, Rose DA, et al. A Public Health Systems View of Risk Communication About Zika. Public Health Rep. 2020 May 1;135(3):343–53.

36. Akhtar Z, Mah-E-Muneer S, Rashid MdM, Ahmed MdS, Islam MdA, Chowdhury S, et al. Antibiotics Use and Its Knowledge in the Community: A Mobile Phone Survey during the COVID-19 Pandemic in Bangladesh. Antibiotics. 2021 Aug 29;10(9):1052.

37. Assefa N, Sié A, Wang D, Korte ML, Hemler EC, Abdullahi YY, et al. Reported Barriers to Healthcare Access and Service Disruptions Caused by COVID-19 in Burkina Faso, Ethiopia, and Nigeria: A Telephone Survey. Am J Trop Med Hyg. 2021 Aug 11;105(2):323–30.

38. Boehm K, Ziewers S, Brandt MP, Sparwasser P, Haack M, Willems F, et al. Telemedicine Online Visits in Urology During the COVID-19 Pandemic—Potential, Risk Factors, and Patients’ Perspective. Eur Urol. 2020 Jul 1;78(1):16–20.

39. Bonet-Esteve A, Muñoz-Miralles R, Gonzalez-Claramunt C, Rufas AM, Cruz XP, Vidal-Alaball J. Influenza vaccination during the coronavirus pandemic: intention to vaccinate among the at-risk population in the Central Catalonia Health Region (VAGCOVID). BMC Fam Pract. 2021 Dec;22(1):84.

40. Bennett D, De Vita E, Ventura V, Bernazzali S, Fossi A, Paladini P, et al. Impact of SARS□CoV□2 outbreak on heart and lung transplant: A patient□perspective survey. Transpl Infect Dis. 2020 Aug 10;e13428.

41. Boscolo-Rizzo P, Guida F, Polesel J, Marcuzzo AV, Capriotti V, D’Alessandro A, et al. Sequelae in adults at 12 months after mild-to-moderate coronavirus disease 2019 (COVID-19). Int Forum Allergy Rhinol. 2021;11(12):1685–8.

42. Chan EYY, Lo ESK, Huang Z, Lam HCY, Yeung MP shan, Kwok K on, et al. Sociodemographic predictors of knowledge, mosquito bite patterns and protective behaviors concerning vector borne disease: The case of dengue fever in Chinese subtropical city, Hong Kong. Beechler BR, editor. PLoS Negl Trop Dis. 2021 Jan 19;15(1):e0008993.

43. Checcucci E, De Luca S, Alessio P, Verri P, Granato S, De Cillis S, et al. Implementing telemedicine for the management of benign urologic conditions: a single centre experience in Italy. World J Urol. 2021 Aug 1;39(8):3109–15.

44. Chen F, Zang Y, Liu Y, Wang X, Lin X. Dispatched nurses’ experience of wearing full gear personal protective equipment to care for COVID□19 patients in China—A descriptive qualitative study. J Clin Nurs. 2021 Jul;30(13–14):2001–14.

45. Cheng FS, Yen YF, Lin SY, Weng SH, Chou YC, Chu D, et al. Prevalence and Factors Associated with the Reuse of Mask during the COVID-19 Pandemic: A Nationwide Survey in Taiwan. Int J Environ Res Public Health. 2021 Jul 29;18(15):8065.

46. Cowling BJ, Ng DMW, Ip DKM, Liao Q, Lam WWT, Wu JT, et al. Community Psychological and Behavioral Responses through the First Wave of the 2009 Influenza A(H1N1) Pandemic in Hong Kong. J Infect Dis. 2010 Sep 15;202(6):867–76.

47. Earle-Richardson G, Prue C, Turay K, Thomas D. Influences of Community Interventions on Zika Prevention Behaviors of Pregnant Women, Puerto Rico, July 2016–June 20171. Emerg Infect Dis. 2018 Dec;24(12):2251–61.

48. Ferrarotti I, Ottaviani S, Balderacchi AM, Barzon V, Silvestri AD, Piloni D, et al. COVID-19 infection in severe Alpha 1-antitrypsin deficiency: Looking for a rationale. Respir Med. 2021 Jul 1;183.

49. Fritzell C, Raude J, Kazanji M, Flamand C. Emerging trends of Zika apprehension in an epidemic setting. PLoS Negl Trop Dis. 2018 Jan 25;12(1):e0006167.

50. Goodwin R, Sun S. Early responses to H7N9 in southern Mainland China. BMC Infect Dis. 2014 Dec;14(1):8.

51. Harling G, Gómez-Olivé FX, Tlouyamma J, Mutevedzi T, Kabudula CW, Mahlako R, et al. Protective Behaviors and Secondary Harms Resulting From Nonpharmaceutical Interventions During the COVID-19 Epidemic in South Africa: Multisite, Prospective Longitudinal Study. JMIR Public Health Surveill. 2021 May 13;7(5):e26073.

52. Heo JY, Chang SH, Go MJ, Kim YM, Gu SH, Chun BC. Risk Perception, Preventive Behaviors, and Vaccination Coverage in the Korean Population during the 2009–2010 Pandemic Influenza A (H1N1): Comparison between High-Risk Group and Non–High-Risk Group. PLoS ONE. 2013 May 17;8(5):e64230.

53. Jayakody S, Hewage SA, Wickramasinghe ND, Piyumanthi RAP, Wijewickrama A, Gunewardena NS, et al. ‘Why are you not dead yet?’ – dimensions and the main driving forces of stigma and discrimination among COVID-19 patients in Sri Lanka. Public Health. 2021 Oct 1;199:10–6.

54. Jones TF, Amanda IL, Craig AS, Schaffner W. Determinants of Influenza Vaccination, 2003–2004: Shortages, Fallacies and Disparities. Clin Infect Dis. 2004 Dec 15;39(12):1824–8.

55. Kirkegaard P, Edwards A, Andersen B. Balancing risks: Qualitative study of attitudes, motivations and intentions about attending for mammography during the COVID-19 pandemic. Scand J Public Health. 2021 Nov;49(7):700–6.

56. Kiviniemi MT, Ram PK, Kozlowski LT, Smith KM. Perceptions of and willingness to engage in public health precautions to prevent 2009 H1N1 influenza transmission. BMC Public Health. 2011 Dec;11(1):152.

57. Lau JTF, Yeung NCY, Choi KC, Cheng MYM, Tsui HY, Griffiths S. Acceptability of A/H1N1 vaccination during pandemic phase of influenza A/H1N1 in Hong Kong: population based cross sectional survey. BMJ. 2009 Oct 27;339:b4164.

58. Lau JT, Griffiths S, Choi KC, Tsui HY. Avoidance behaviors and negative psychological responses in the general population in the initial stage of the H1N1 pandemic in Hong Kong. BMC Infect Dis. 2010 May 28;10(1):139.

59. Lau JTF, Griffiths S, Choi K chow, Lin C. Prevalence of preventive behaviors and associated factors during early phase of the H1N1 influenza epidemic. Am J Infect Control. 2010 Jun;38(5):374–80.

60. Lau J, Yang X, Tsui H, Kim J. Monitoring community responses to the SARS epidemic in Hong Kong: from day 10 to day 62. J Epidemiol Community Health. 2003 Nov;57(11):864–70.

61. Lau JTF, Yang X, Pang E, Tsui HY, Wong E, Wing YK. SARS-related Perceptions in Hong Kong. Emerg Infect Dis. 2005 Mar;11(3):417–24.

62. Parsons Leigh J, Kemp LG, de Grood C, Brundin-Mather R, Stelfox HT, Ng-Kamstra JS, et al. A qualitative study of physician perceptions and experiences of caring for critically ill patients in the context of resource strain during the first wave of the COVID-19 pandemic. BMC Health Serv Res. 2021 Dec;21(1):374.

63. Liao Q, Cowling B, Lam WT, Ng MW, Fielding R. Situational Awareness and Health Protective Responses to Pandemic Influenza A (H1N1) in Hong Kong: A Cross-Sectional Study. Ng LFP, editor. PLoS ONE. 2010 Oct 12;5(10):e13350.

64. Liao Q, Cowling BJ, Lam WW, Ng DM, Fielding R. Anxiety, worry and cognitive risk estimate in relation to protective behaviors during the 2009 influenza A/H1N1 pandemic in Hong Kong: ten cross-sectional surveys. BMC Infect Dis. 2014 Mar 27;14:169.

65. Liao Q, Xiao J, Cheung J, Ng TWY, Lam WWT, Ni MY, et al. Community psychological and behavioural responses to coronavirus disease 2019 over one year of the pandemic in 2020 in Hong Kong. Sci Rep. 2021 Dec;11(1):22480.

66. Lin Y, Huang L, Nie S, Liu Z, Yu H, Yan W, et al. Knowledge, Attitudes and Practices (KAP) related to the Pandemic (H1N1) 2009 among Chinese General Population: a Telephone Survey. BMC Infect Dis. 2011 Dec;11(1):128.

67. Liu Q, Luo D, Haase JE, Guo Q, Wang XQ, Liu S, et al. The experiences of health-care providers during the COVID-19 crisis in China: a qualitative study. Lancet Glob Health. 2020 Jun 1;8(6):e790–8.

68. Lupton D, Lewis S. Learning about COVID-19: a qualitative interview study of Australians’ use of information sources. BMC Public Health. 2021 Dec;21(1):662.

69. Markkanen P, Brouillette N, Quinn M, Galligan C, Sama S, Lindberg J, et al. “It changed everything”: The Safe Home Care qualitative study of the COVID-19 pandemic’s impact on home care aides, clients, and managers. BMC Health Serv Res. 2021 Oct 5;21(1):1055.

70. Mayeur A, Binois O, Gallot V, Hesters L, Benoit A, Oppenheimer A, et al. First follow-up of art pregnancies in the context of the COVID-19 outbreak. Eur J Obstet Gynecol Reprod Biol. 2020 Oct;253:71–5.

71. Meng H, Liao Q, Suen LKP, O’Donoghue M, Wong CM, Yang L. Healthcare seeking behavior of patients with influenza like illness: comparison of the summer and winter influenza epidemics. BMC Infect Dis. 2016 Sep 20;16(1):499.

72. Mora AM, Lewnard JA, Rauch S, Kogut K, Jewell N, Cuevas M, et al. Impact of COVID-19 Pandemic on California Farmworkers’ Mental Health and Food Security. J Agromedicine. 2022 Apr 11;1–12.

73. Qian M, Wu Q, Wu P, Hou Z, Liang Y, Cowling BJ, et al. Anxiety levels, precautionary behaviours and public perceptions during the early phase of the COVID-19 outbreak in China: a population-based cross-sectional survey. BMJ Open. 2020 Oct 8;10(10):e040910.

74. Quah SR, Hin-Peng L. Crisis Prevention and Management during SARS Outbreak, Singapore. Emerg Infect Dis. 2004;10(2):364–8.

75. Raude J, Peretti-Watel P, Ward J, Flamand C, Verger P. Are Perceived Prevalences of Infection also Biased and How? Lessons from Large Epidemics of Mosquito-Borne Diseases in Tropical Regions. Med Decis Making. 2018 Apr;38(3):377–89.

76. Reed C, Angulo FJ, Biggerstaff M, Swerdlow D, Finelli L. Influenza-Like Illness in the Community during the Emergence of 2009 Pandemic Influenza A(H1N1) – Survey of 10 States, April 2009. Clin Infect Dis. 2011 Jan 1;52(suppl_1):S90–3.

77. Shah A, Guessi M, Wali S, Ware P, McDonald M, O’Sullivan M, et al. The Resilience of Cardiac Care Through Virtualized Services During the COVID-19 Pandemic: Case Study of a Heart Function Clinic. JMIR Cardio. 2021 May 5;5(1):e25277.

78. Shati M, Alami A, Mortazavi SS, Eybpoosh S, Emamian MH, Moghadam M. Adherence to Self-isolation measures by older adults during coronavirus disease 2019 (COVID-19) epidemic: A phone survey in Iran. Med J Islam Repub Iran. 2020 Nov 10;34:152.

79. SteelFisher GK, McMurtry CL, Caporello HL, McGowan E, Schafer TJ, Lubell KM, et al. Experiences and Views of Domestic Summer Travelers During the COVID-19 Pandemic: Findings from a National Survey. Health Secur. 2021 Jun 1;19(3):338–48.

80. Taglioni F, Cartoux M, Dellagi K, Dalban C, Fianu A, Carrat F, et al. The influenza A (H1N1) pandemic in Reunion Island: knowledge, perceived risk and precautionary behaviour. BMC Infect Dis. 2013 Jan 24;13:34.

81. Tan EK, Koh YX, Kee T, Juhari JB, Tan TE, Sim DKL, et al. Waitlisted Transplant Candidates’ Attitudes and Concerns Toward Transplantation During COVID-19. Ann Transplant. 2020 Dec 8;25:e926992.

82. Topcu S, Kilic Ucar A, Güven B, Akyüz A. Investigation of health care needs, behaviors, and existing health conditions of individuals aged ≥ 65 during the pandemic. Educ Gerontol. 2022 Feb 1;48(2):64–73.

83. Wong LP, Sam IC. Public Sources of Information and Information Needs for Pandemic Influenza A(H1N1). J Community Health. 2010 Dec 1;35(6):676–82.

84. Wong LP, Sam IC. Temporal changes in psychobehavioral responses during the 2009 H1N1 influenza pandemic. Prev Med. 2010 Jul;51(1):92–3.

85. Wong LP, Sam IC. Behavioral responses to the influenza A(H1N1) outbreak in Malaysia. J Behav Med. 2011;34(1):23–31.

86. Wong LP, Sam IC. Knowledge and Attitudes in Regard to Pandemic Influenza A(H1N1) in a Multiethnic Community of Malaysia. Int J Behav Med. 2011;18(2):112–21.

87. Xiao J, Cheung JK, Wu P, Ni MY, Cowling BJ, Liao Q. Temporal changes in factors associated with COVID-19 vaccine hesitancy and uptake among adults in Hong Kong: Serial cross-sectional surveys. Lancet Reg Health – West Pac [Internet]. 2022 Jun 1 [cited 2022 Jun 16];23. Available from: https://www.thelancet.com/journals/lanwpc/article/PIIS2666-6065(22)00056-6/fulltext

88. Xin M, Lau JT fai, Lau MMC. Multi-dimensional factors related to participation in a population-wide mass COVID-19 testing program among Hong Kong adults: A population-based randomized survey. Soc Sci Med 1982. 2022 Feb;294:114692.

89. Yeung NCY, Lau JTF, Choi KC, Griffiths S. Population Responses during the Pandemic Phase of the Influenza A(H1N1)pdm09 Epidemic, Hong Kong, China. Emerg Infect Dis. 2017 May;23(5):813–5.

90. Yin Y, Chu X, Han X, Cao Y, Di H, Zhang Y, et al. General practitioner trainees’ career perspectives after COVID-19: a qualitative study in China. BMC Fam Pract. 2021 Dec;22(1):18.

