## Supplemental Materials for "Use of Interviewer-Administered Telephone Surveys during Infectious Disease Outbreaks, Epidemics, and Pandemics: A Scoping Review"

**Supplemental Material 1** List of Included Studies

| Study ID | Author (Year) | Journal | Origin | Sample Size | Design | Scale | Population | Infectious Disease | Phone Type |
|---|---|---|---|---|---|---|---|---|---|
| #1 | Abba et al[14] (2021) | Afrika Focus | Nigeria | 11 | Qualitative | Local | HCPs | COVID-19 | Mobile |
| #2 | Abdelmagid et al[31] (2021) | BMC Public Health | Sudan | 89 | Qualitative | Regional | Other: Adult Individuals, Patients | COVID-19 | Not Specified |
| #3 | Akhtar et al[36] (2021) | Antibiotics | Banglade sh | 1845 | Quantitative Descriptive | National | Adult Individuals | COVID-19 | Mobile |
| #4 | Assefa et al[37] (2021) | American Society of Tropical Medicine and Hygiene | Burkina Faso, Ethiopia, Nigeria | 10797 | Quantitative Randomized Controlled Trials | Internatio nal | Other: Adult Individuals, HCPs | COVID-19 | Not Specified |
| #5 | Boehm et al[38] (2020) | European Urology | Germany | 399 | Quantitative Non-Randomized | Local | Patients | COVID-19 | Not Specified |
| #6 | Bonet-Esteve et al[39] (2021) | BMC Family Practice | Spain | 434 | Quantitative Non-Randomized | Regional | Patients | COVID-19 | Not Specified |
| #7 | Bonnett et al[40] (2021) | Transplant Infectious Disease | Italy | 144 | Quantitative Non-Randomized | Local | Patients | COVID-19 | Not Specified |
| #8 | Boscolo-Rizzo et al[41] (2021) | International Forum of Allergy & Rhinology | Italy | 304 | Quantitative Descriptive | Regional | Patients | COVID-19 | Not Specified |
| #9 | Chan et al[42] (2021) | PLOS Neglected Tropical Diseases | China | 590 | Quantitative Descriptive | Local | Adult Individuals | Dengue | Landline |
| #10 | Checcucci et al[43] (2021) | World Journal of Urology | Italy | 607 | Quantitative Descriptive | Local | Patients | COVID-19 | Not Specified |

| | | | | | | | | | |
|---|---|---|---|---|---|---|---|---|---|
| #11 | Chen et al[44] (2021) | Journal of Clinical Nursing | China | 15 | Qualitative | Local | HCPs | COVID-19 | Not Specified |
| #12 | Cheng et al[45] (2021) | International Journal of Environmental Research and Public Health | China | 1075 | Quantitative Descriptive | Local | Adult Individuals | COVID-19 | Not Specified |
| #13 | Cowling et al[46] (2010) | Journal of Infectious Diseases | China | 12965 | Quantitative Descriptive | Local | Adult Individuals | H1N1 | Landline |
| #14 | Earle-Richardson et al[47] (2018) | Emerging Infectious Diseases | United States | 950 | Quantitative Non-Randomized | Regional | Other: Women | Zika | Not Specified |
| #15 | Ferrarotti et al[48] (2021) | Respiratory Medicine | Italy | 209 | Quantitative Descriptive | National | Patients | COVID-19 | Not Specified |
| #16 | Fritzell et al[49] (2018) | PLOS Neglected Tropical Diseases | France | 1129 | Quantitative Non-Randomized | Regional | Adult Individuals | Zika | 50-50 |
| #17 | Gao et al[15] (2021) | BMJ Open | China | 31332 | Quantitative Non-Randomized | Local | Adult Individuals | COVID-19 | Not Specified |
| #18 | Goodwin et al[50] (2014) | BMC Infectious Diseases | China | 1011 | Quantitative Descriptive | Local | Adult Individuals | H7N9 | Landline |
| #19 | Harling et al[51] (2021) | JMIR Public Health and Surveillance | South Africa | 5120 | Quantitative Non-Randomized | Regional | Households | COVID-19 | Mobile |
| #20 | Heo et al[52] (2013) | PLOS One | South Korea | 1650 | Quantitative Non-Randomized | National | Adult Individuals | H1N1 | Not Specified |
| #21 | Jayakody et al[53] (2021) | Public Health | Sri Lanka | 139 | Qualitative | Regional | Patients | COVID-19 | Not Specified |

| | | | | | | | | | |
|---|---|---|---|---|---|---|---|---|---|
| #22 | Jones et al[54] (2004) | Clinical Infectious Diseases | United States | 4028 | Quantitative Non-Randomized | Local | Adult Individuals | Seasonal Flu | Landline |
| #23 | Kirkegaard et al[55] (2021) | Scandinavian Journal of Public Health | Denmark | 33 | Qualitative | Regional | Other: Women | COVID-19 | Not Specified |
| #24 | Kiviniemi et al[56] (2011) | BMC Public Health | United States | 807 | Quantitative Descriptive | Local | Adult Individuals | H1N1 | Not Specified |
| #25 | Kuehne et al[23] (2016) | PLOS Neglected Tropical Diseases | Liberia | 905 | Quantitative Descriptive | Regional | Households | Ebola | Mobile |
| #26 | Lau et al[57] (2009) | British Medical Journal | China | 301 | Quantitative Descriptive | Local | Adult Individuals | H1N1 | Landline |
| #27 | Lau et al[58] (2010) | BMC Infectious Diseases | China | 999 | Quantitative Descriptive | Local | Adult Individuals | H1N1 | Landline |
| #28 | Lau et al[59] (2010) | American Journal of Infection Control | China | 999 | Quantitative Descriptive | Local | Adult Individuals | H1N1 | Landline |
| #29 | Lau et al[60] (2003) | Journal of Epidemiology and Community Health | China | 1397 | Quantitative Descriptive | Local | Adult Individuals | SARS | Landline |
| #30 | Lau et al[61] (2005) | Emerging Infectious Diseases | China | 1681 | Quantitative Descriptive | Local | Adult Individuals | SARS | Landline |
| #31 | Lee et al[32] (2020) | International Journal of Environmental Research and Public Health | South Korea | 18 | Qualitative | Local | HCPs | COVID-19 | Mobile |
| #32 | Leigh et al[62] (2021) | BMC Health Services Research | Canada | 15 | Qualitative | Regional | HCPs | COVID-19 | Not Specified |

| | | | | | | | | | |
|---|---|---|---|---|---|---|---|---|---|
| #33 | Liao et al[63] (2010) | PLOS One | China | 1001 | Quantitative Non-Randomized | Local | Adult Individuals | H1N1 | Landline |
| #34 | Liao et al[17] (2011) | International Journal of Behavioral Medicine | China | 2776 | Quantitative Non-Randomized | Local | Households | H1N1, H5N1 | Landline |
| #35 | Liao et al[64] (2014) | BMC Infectious Diseases | China | 10345 | Quantitative Non-Randomized | Local | Households | H1N1 | Landline |
| #36 | Liao et al[65] (2021) | Scientific Reports | China | 30827 | Quantitative Non-Randomized | Local | Adult Individuals | COVID-19 | 50-50 |
| #37 | Lin et al[66] (2011) | BMC Infectious Diseases | China | 10669 | Quantitative Descriptive | Regional | Adult Individuals | H1N1 | Landline |
| #38 | Liu et al[67] (2020) | Lancet Global Health | China | 13 | Qualitative | Local | HCPs | COVID-19 | Not Specified |
| #39 | Lupton et al[68] (2021) | BMC Public Health | Australia | 40 | Qualitative | National | Adult Individuals | COVID-19 | Not Specified |
| #40 | Markkanen et al[69] (2021) | BMC Health Services Research | United States | 37 | Qualitative | Local | Other: Elderly, Home Care Workers | COVID-19 | Not Specified |
| #41 | Mayeur et al[70] (2020) | European Journal of Obstetrics Gynecology and Reproductive Biology | France | 104 | Quantitative Descriptive | Local | Other: Women | COVID-19 | Not Specified |
| #42 | Meng et al[71] (2016) | BMC Infectious Diseases | China | 516 | Quantitative Non-Randomized | Local | Households | Seasonal Flu | Landline |
| #43 | Mora et al[72] (2022) | Journal of Agromedicine | United States | 1115 | Quantitative descriptive | Local | Other: Farm Workers | COVID-19 | Mobile |

| | | | | | | | | | |
|---|---|---|---|---|---|---|---|---|---|
| #44 | Mwamba et al[33] (2020) | Public Health Action | Zambia | 17 | Qualitative | Local | Patients | COVID-19 | Mobile |
| #45 | Qian et al[73] (2020) | BMJ Open | China | 1011 | Quantitative Non-Randomized | Regional | Adult Individuals | COVID-19 | Landline |
| #46 | Quah et al[74] (2004) | Emerging Infectious Diseases | Singapor e | 1202 | Quantitative descriptive | National | Adult Individuals | SARS | Not Specified |
| #47 | Raude et al[75] (2018) | Medical Decision Making | France | 3721 | Quantitative descriptive | Regional | Adult Individuals | Chikungunya, Dengue, Zika | Not Specified |
| #48 | Reed et al[76] (2011) | Clinical Infectious Diseases | United States | 1790 | Quantitative Descriptive | Regional | Households | H1N1 | Not Specified |
| #49 | Rubin et al[34] (2016) | European Journal of Psycho traumatology | United Kingdom | 51 | Qualitative | Regional | Other: Public Health Professional s, NGO, University Researcher | Ebola | Not Specified |
| #50 | Sell et al[35] (2020) | Public Health Reports | United States | 28 | Qualitative | Regional | Other: Public Health Professionals | Zika | Not Specified |
| #51 | Shah et al[77] (2021) | JMIR Cardio | Canada | 29 | Qualitative | Local | Other: HCPs, Patients | COVID-19 | Not Specified |
| #52 | Shati et al[78] (2020) | Medical Journal of The Islamic Republic of Iran | Iran | 558 | Quantitative Descriptive | National | Other: Elderly | COVID-19 | Not Specified |
| #53 | Smaghi et al[24] (2021) | International Journal of Infectious Diseases | Papua New Guinea | 426 | Mixed Methods | National | HCPs | COVID-19 | Not Specified |
| #54 | SteelFisher et al[79] (2021) | Health Security | United States | 1986 | Quantitative Non-Randomized | National | Adult Individuals | COVID-19 | Mobile |

| | | | | | | | | | |
|---|---|---|---|---|---|---|---|---|---|
| #55 | Stoop et al[16] (2021) | World Development | Democratic Republic of Congo | 1000 | Quantitative Non-Randomized | Regional | Other: Households, Small Businesses | Covid-19, Ebola | Not Specified |
| #56 | Taglioni et al[80] (2013) | BMC Infectious Diseases | France | 725 | Quantitative Descriptive | Regional | Households | H1N1 | Not Specified |
| #57 | Tan et al[81] (2020) | Annals of Transplantation | Singapore | 71 | Quantitative Descriptive | Local | Patients | COVID-19 | Not Specified |
| #58 | Topcu et al[82] (2022) | Educational Gerontology | Turkey | 134 | Quantitative Descriptive | Regional | Patients | COVID-19 | Not Specified |
| #59 | Wong et al[83] (2010) | Journal of Community Health | Malaysia | 1050 | Quantitative Descriptive | Regional | Adult Individuals | H1N1 | Landline |
| #60 | Wong et al[84] (2010) | Preventive Medicine | Malaysia | 1050 | Quantitative Descriptive | Local | Adult Individuals | H1N1 | Landline |
| #61 | Wong et al[85] (2011) | Journal of Behavioral Medicine | Malaysia | 1050 | Quantitative Descriptive | Local | Adult Individuals | H1N1 | Landline |
| #62 | Wong et al[86] (2011) | International Journal of Behavioral Medicine | Malaysia | 1050 | Quantitative Descriptive | Regional | Adult Individuals | H1N1 | Landline |
| #63 | Xiao et al[87] (2022) | Lancet Regional Health Western Pacific | China | 7411 | Quantitative Descriptive | Local | Adult Individuals | COVID-19 | 50-50 |
| #64 | Xin et al[88] (2022) | Social Science & Medicine | China | 443 | Quantitative Descriptive | Local | Adult Individuals | COVID-19 | Landline |
| #65 | Yeung et al[89] (2017) | Emerging Infectious Diseases | China | 503 | Quantitative Descriptive | Local | Adult Individuals | H1N1 | Landline |

| | | | | | | | | | |
|---|---|---|---|---|---|---|---|---|---|
| #66 | Yin et al[90] (2021) | BMC Family Practice | China | 12 | Qualitative | Regional | HCPs | COVID-19 | Not Specified |
| #67* | Ba et al[28] (2022) | Human Vaccines & Immuno therapeutics | Senegal | 637 | Mixed Methods | National | Adult Individuals | COVID-19 | Mobile |
| #68* | Diallo et al[20] (2022) | Revue d'Épidémiologie et de Santé Publique | Senegal | 813 | Quantitative Descriptive | National | Adult Individuals | COVID-19 | Not Specified |
| #69* | Ridde et al[21] (2021) | Human Vaccines & Immuno therapeutics | Senegal | 607 | Quantitative Descriptive | National | Adult Individuals | COVID-19 | Mobile |
| #70* | Ridde et al[22] (2022) | PLOS Global Public Health | Senegal | 843 | Mixed Methods | National | Adult Individuals | COVID-19 | Mobile |

*These studies were manually added to supplement the database searches.

**Supplemental Material 2** List of Included Studies (Continued)

| Study ID | Title | Purpose |
|---|---|---|
| #1 | Experiences of Healthcare Workers of Hospitalised Individuals with COVID-19 in Kano, Nigeria: A Qualitative Study | to explore the experiences of healthcare workers managing hospitalised patients with covid-19 in a treatment centre in Kano, Nigeria |
| #2 | Acceptability and feasibility of strategies to shield the vulnerable during the COVID-19 outbreak: a qualitative study in six Sudanese communities | to explore the acceptability and feasibility of strategies to shield persons at higher risk of severe COVID-19 outcomes, during the COVID-19 epidemic in six communities in Sudan |
| #3 | Antibiotics Use and its Knowledge in the Community: A Mobile Phone Survey during the COVID-19 Pandemic in Bangladesh | to assess antibiotic use for any reported illnesses in the preceding four weeks and knowledge regarding antibiotics among the general population in Bangladesh |
| #4 | Reported Barriers to Healthcare Access and Service Disruptions Caused by COVID-19 in Burkina Faso, Ethiopia, and Nigeria: A Telephone Survey | to collect data regarding the effects of COVID-19 on the healthcare system from the perspectives of two groups of stakeholders: healthcare providers and community members |
| #5 | Telemedicine Online Visits in Urology During the COVID-19 Pandemic-Potential, Risk Factors, and Patients' Perspective | to evaluate patients’ eligibility for telemedicine according to the physician and examined the patients’ perspective by evaluating their willingness for telemedicine |

| | | |
|---|---|---|
| #6 | Influenza Vaccination during the Coronavirus Pandemic: Intention to Vaccinate among the At-Risk Population in the Central Catalonia Health Region (VAGCOVID) | to determine the at-risk population’s intention to vaccinate against seasonal influenza during the 2020-21 flu campaign in the Central Catalonia Health Region, in the midst of the SARS-CoV-2 pandemic |
| #7 | Impact of SARS-CoV-2 Outbreak on Heart and Lung Transplant: A Patient-Perspective Survey | to evaluate the incidence of COVID-19 and the impact of the SARS-CoV-2 outbreak on the personal hygiene and expectations in heart and lung transplant recipients |
| #8 | Sequelae in Adults at 12 Months after Mild-to-Moderate Coronavirus Disease 2019 (COVID-19) | to evaluate the prevalence of COVID-related symptoms 12-months after the onset of mild-to-moderate disease |
| #9 | Sociodemographic Predictors of Knowledge, Mosquito Bite Patterns and Protective Behaviors Concerning Vector Borne Disease: The Case of Dengue Fever in Chinese Subtropical City, Hong Kong | to examine the knowledge of dengue, mosquito bite patterns, protective behavior practices and their associated factors |
| #10 | Implementing Telemedicine for the Management of Benign Urologic Conditions: A Single Centre Experience in Italy | to evaluate the health status of these patients, to identify those who needed an “in-person” ambulatory visit due to the worsening of their condition |
| #11 | Dispatched Nurses' Experience of Wearing Full Gear Personal Protective Equipment to Care for COVID-19 Patients in China—A Descriptive Qualitative Study | to explore dispatched nurses’ experiences of wearing full gear personal protective equipment to care for patients with coronavirus disease-2019 (COVID-19) in Wuhan, China |
| #12 | Prevalence and Factors Associated with the Reuse of Mask during the COVID-19 Pandemic: A Nationwide Survey in Taiwan | to understand the factors associated with mask reuse and provide important information for educating individuals regarding appropriate preventive behaviors against the SARS-COV-2 infection |
| #13 | Community Psychological and Behavioral Responses through the First Wave of the 2009 Influenza A(H1N1) Pandemic in Hong Kong | to examine trends in anxiety, risk perception, knowledge on modes of transmission, and preventive behaviors |
| #14 | Influences of Community Interventions on Zika Prevention Behaviors of Pregnant Women, Puerto Rico, July 2016–June 2017 | to assess how community education efforts influenced pregnant women’s Zika prevention behaviors during the 2016 Centers for Disease Control and Prevention–Puerto Rico Department of Health Zika virus response |
| #15 | COVID-19 Infection in Severe Alpha 1-Antitrypsin Deficiency: Looking for a Rationale | to investigate whether people with severe AAT deficiency (AATD) have an increased risk of (severe) COVID-19 infection |
| #16 | Emerging Trends of Zika Apprehension in an Epidemic Setting | to examine public perceptions associated with this new health threat, with the purpose of informing ongoing intervention practices |
| #17 | Pandemic fatigue and attenuated impact of avoidance behaviours against COVID-19 transmission in Hong Kong by cross-sectional telephone surveys | to explore the attenuated impact of reported avoidance behaviours adherence on the transmission of COVID-19 |
| #18 | Early Responses to H7N9 in Southern Mainland China | to examine risk awareness and media use, beliefs about the emergence of the threat and those most at risk, anxiety about infection and preventive and avoidant behaviours |

| | | |
|---|---|---|
| #19 | Protective Behaviors and Secondary Harms Resulting From Nonpharmaceutical Interventions During the COVID-19 Epidemic in South Africa: Multisite, Prospective Longitudinal Study | to observe how households in rural and peri-urban areas responded to, and were affected by, national NPIs enacted to minimize the epidemic spread of Covid-19 |
| #20 | Risk Perception, Preventive Behaviors, and Vaccination Coverage in the Korean Population during the 2009–2010 Pandemic Influenza A (H1N1): Comparison between High-Risk Group and Non–High-Risk Group | to estimate the vaccination coverage, public perception, and preventive behaviors against pandemic influenza A (H1N1) and to understand the motivation and barriers to vaccination between high-risk and non–high-risk groups during the outbreak of pandemic influenza A (H1N1) |
| #21 | Why Are You Not Dead Yet?' - Dimensions and the Main Driving Forces of Stigma and Discrimination among COVID-19 Patients in Sri Lanka | to explore the experiences, and main driving forces of stigma and discrimination among COVID-19 patients, following hospital discharge, in Sri Lanka |
| #22 | Determinants of Influenza Vaccination, 2003--2004: Shortages, Fallacies and Disparities | to assess people's knowledge, attitudes, and beliefs about vaccination and to assess how access issues may have impacted other determinants of vaccination rates |
| #23 | Balancing Risks: Qualitative Study of Attitudes, Motivations and Intentions about Attending for Mammography during the COVID-19 Pandemic | to explore the attitudes, motivations and intentions around attending for mammography among women in the Danish population-based breast cancer screening programme who cancelled or postponed mammography during, and due to, the COVID-19 pandemic |
| #24 | Perceptions of and willingness to engage in public health precautions to prevent 2009 H1N1 influenza transmission | to examine individuals' interpretation of recommendations, willingness to comply, and factors predicting willingness |
| #25 | Mortality, Morbidity and Health-Seeking Behaviour during the Ebola Epidemic 2014–2015 in Monrovia Results from a Mobile Phone Survey | to quantify mortality and morbidity and describe health-seeking behaviour in Monrovia |
| #26 | Acceptability of A/H1N1 Vaccination during Pandemic Phase of Influenza A/H1N1 in Hong Kong: Population Based Cross Sectional Survey | to investigate the intention of the Hong Kong general population to take up vaccination against influenza A/H1N1 |
| #27 | Avoidance Behaviors and Negative Psychological Responses in the General Population in the Initial Stage of the H1N1 Pandemic in Hong Kong | to examine the prevalence of the avoidance behaviors (i.e. avoiding going out, visiting crowded places and visiting hospitals) and negative psychological responses of the general population in Hong Kong at the initial stage of the H1N1 epidemic |
| #28 | Prevalence of preventive behaviors and associated factors during early phase of the H1N1 influenza epidemic | to investigate the prevalence of self-reported preventive behaviors in response to the influenza A/H1N1epidemic in Hong Kong, including wearing face masks regularly in public areas, wearing face masks in case of influenza-like illness (ILI) symptoms, and frequent handwashing |
| #29 | Monitoring community responses to the SARS epidemic in Hong Kong: from day 10 to day 62 | to report the evolution in perceptions and behaviours of the general public in response to the severe acute respiratory syndrome (SARS) epidemic in Hong Kong |

| | | |
|---|---|---|
| #30 | SARS-related Perceptions in Hong Kong | to understand different aspects of community responses related to severe acute respiratory syndrome (SARS) |
| #31 | South Korean Nurses' Experiences with Patient Care at a COVID-19-Designated Hospital: Growth after the Frontline Battle against an Infectious Disease | to explore the lived experiences and essential structure of the experiences of a group of nurses at a COVID-19-designated hospital who were providing patient care |
| #32 | A qualitative study of physician perceptions and experiences of caring for critically ill patients in the context of resource strain during the first wave of the COVID-19 pandemic | to investigate physicians' perceptions and experiences of caring for critically ill patients in the context of actual or anticipated resource strain during the COVID-19 pandemic, and to explore implications for the healthcare workforce and the delivery of patient care |
| #33 | Situational awareness and health protective responses to pandemic influenza A (H1N1) in Hong Kong: a cross-sectional study | to test a hypothesized model of associations between trust in (formal/ informal) information, situational awareness variables (causal understanding, self-efficacy, susceptibility and worry) and different types of health protective behaviours (hand hygiene and social distancing) for influenza protection |
| #34 | The Influence of Social-Cognitive Factors on Personal Hygiene Practices to Protect Against Influenzas: Using Modelling to Compare Avian A/H5N1 and 2009 Pandemic A/ H1N1 Influenzas in Hong Kong | to model associations between trust in information, perceived hygiene effectiveness, knowledge about the causes of influenza, perceived susceptibility and worry, and personal hygiene practices (PHPs) associated with influenza |
| #35 | Anxiety, worry and cognitive risk estimate in relation to protective behaviors during the 2009 influenza A/H1N1 pandemic in Hong Kong: ten cross-sectional surveys | to compare the strength of associations between different cognitive and affective measures of risk and self-reported protective behaviors |
| #36 | Community psychological and behavioural responses to coronavirus disease 2019 over one year of the pandemic in 2020 in Hong Kong | to monitor the changes of public stress appraisal including COVID-19 risk perception, personal efficacy and confidence in government's pandemic control, behavioural coping (i.e., precautionary behaviours) and psychological distress over one year of the pandemic in 2020 in Hong Kong |
| #37 | Knowledge, attitudes and practices (KAP) related to the pandemic (H1N1) 2009 among Chinese general population: a telephone survey | to investigate community responses to A/H1N1 in China; to describe the knowledge, attitudes and practices of A/H1N1 among general population in China and put forward policy recommendations to government in case of future similar conditions |
| #38 | The experiences of health-care providers during the COVID-19 crisis in China: a qualitative study | to describe the experiences of these health-care providers in the early stages of the outbreak |
| #39 | Learning about COVID-19: a qualitative interview study of Australians' use of information sources | to investigate how Australians learnt about the novel coronavirus and COVID-19 and what sources of information they had found most useful and valuable during the early months of the pandemic |
| #40 | "It changed everything": The Safe Home Care qualitative study of the COVID-19 pandemic's impact on home care aides, clients, and managers | to characterize qualitatively the impact of the COVID-19 pandemic on three key HC stakeholders: clients, aides, and agency managers |

| | | |
|---|---|---|
| #41 | First follow-up of art pregnancies in the context of the COVID-19 outbreak | to follow up the monitoring, health and anxiety from women who became pregnant after an embryo transfer or a intrauterine insemination during the COVID-19 epidemic in France |
| #42 | Healthcare seeking behavior of patients with influenza like illness: comparison of the summer and winter influenza epidemics | to compare healthcare seeking behaviors, such as seeking medical consultations in western practitioners or Traditional Chinese Medicine (TCM) and self-medication of ILI patients, between the summer and winter influenza epidemics in Hong Kong |
| #43 | Impact of COVID-19 pandemic on California farmworkers' mental health and food security | to examine the mental health and economic impact of the COVID-19 pandemic on Latino farmworkers in California |
| #44 | Diagnosed with TB in the era of COVID-19: patient perspectives in Zambia | to explore the potential of COVID-19 to further compromise TB care engagement in Zambia |
| #45 | Anxiety levels, precautionary behaviours and public perceptions during the early phase of the COVID-19 outbreak in China: a population-based cross-sectional survey | To investigate psychological and behavioural responses to COVID-19 among the Chinese general population |
| #46 | Crisis prevention and management during SARS outbreak, Singapore | to examine four areas of public reaction to the SARS outbreak in Singapore: preventive practices, perception of self-health, knowledge of SARS, and appraisal of SARS crisis management |
| #47 | Are Perceived Prevalences of Infection also Biased and How? Lessons from Large Epidemics of Mosquito-Borne Diseases in Tropical Regions | to examine the accuracy of judgments of risk frequencies |
| #48 | Influenza-Like Illness in the Community during the Emergence of 2009 Pandemic Influenza A(H1N1) – Survey of 10 States, April 2009 | to better estimate the burden of ILI in the community at the time of the emergence of pH1N1 |
| #49 | How to support staff deploying on overseas humanitarian work: a qualitative analysis of responder views about the 2014/15 West African Ebola outbreak | to understand what factors were stressful or uplifting |
| #50 | A Public Health Systems View of Risk Communication About Zika | to characterize state and local risk communication efforts launched in response to Zika |
| #51 | The Resilience of Cardiac Care Through Virtualized Services During the COVID-19 Pandemic: Case Study of a Heart Function Clinic | to understand people’s experiences with and the barriers and facilitators of the rapid virtualization and expansion of cardiac care resulting from the pandemic |
| #52 | Adherence to Self-isolation measures by older adults during coronavirus disease 2019 (COVID-19) epidemic: A phone survey in Iran | to identify the coverage, efficacy, and integrity of self-isolation and its predictors in the Iranian older adults (≥60 years) from February 19 to 19 March 2020 |

| | | |
|---|---|---|
| #53 | Barriers and enablers experienced by health care workers in swabbing for COVID-19 in Papua New Guinea: A multi-methods cross-sectional study | to identify the barriers and enablers that Health Care Workers (HCWs) in Papua New Guinea experienced in swabbing for COVID-19 |
| #54 | Experiences and Views of Domestic Summer Travelers During the COVID-19 Pandemic: Findings from a National Survey | to fill gaps in the scientific understanding of domestic travel-related behaviors during the COVID-19 pandemic by using data from a nationally representative survey of adults in the United States who traveled during the summer of 2020 |
| #55 | Covid-19 vs. Ebola: Impact on households and small businesses in North Kivu, Democratic Republic of Congo | to compare the socioeconomic impact across two high-profile disease outbreaks that affected North Kivu simultaneously |
| #56 | The influenza A (H1N1) pandemic in Reunion Island: knowledge, perceived risk and precautionary behaviour | to investigate the perceived severity, vulnerability and precautionary behaviour adopted in response to the influenza A (H1N1) epidemic that broke out in 2009 on Reunion Island (Indian Ocean) |
| #57 | Waitlisted Transplant Candidates' Attitudes and Concerns Toward Transplantation During COVID-19 | to determine the opinions and concerns of waitlisted candidates for transplant during the current COVID-19 pandemic |
| #58 | Investigation of health care needs, behaviors, and existing health conditions of individuals aged ≥ 65 during the pandemic | to determine the investigation of health-care needs, behaviors, and existing health conditions of individuals aged and over the age of 65 during the pandemic |
| #59 | Public Sources of Information and Information Needs for Pandemic Influenza A(H1N1) | to explore sources of influenza A(H1N1)-related information, specific information needs and preferences of the lay public during the peak of the outbreak |
| #60 | Temporal changes in psychobehavioral responses during the 2009 H1N1 influenza pandemic | to examine the temporal changes in psychobehavioral responses in relation to reported 2009 H1N1 influenza deaths |
| #61 | Behavioral responses to the influenza A(H1N1) outbreak in Malaysia | to assess: (1) fear of the A(H1N1) pandemic; (2) risk avoidance behavior; (3) health-protective behavior; and (4) psychosocial impact in the ethnically diverse population of Malaysia |
| #62 | Knowledge and Attitudes in Regard to Pandemic Influenza A(H1N1) in a Multiethnic Community of Malaysia | to investigate the level of knowledge and attitudes towards the influenza A(H1N1) outbreak across various ethnic groups and socio-demographic backgrounds in Malaysia |
| #63 | Temporal changes in factors associated with COVID-19 vaccine hesitancy and uptake among adults in Hong Kong: Serial cross-sectional surveys | to explore the factors associated with vaccine hesitancy and uptake among adults before and after the implementation of the COVID-19 vaccination program in Hong Kong |
| #64 | Multi-dimensional factors related to participation in a population-wide mass COVID-19 testing program among Hong Kong adults: A population-based randomized survey | to investigate the multi-dimensional factors associated with participation in a free and voluntary population-wide mass COVID-19 testing program |

| | | |
|---|---|---|
| #65 | Population Responses during the Pandemic Phase of the Influenza A(H1N1)pdm09 Epidemic, Hong Kong, China | to explore changes in their behavioral and psychological responses to the influenza A(H1N1)pdm09 virus epidemic |
| #66 | General practitioner trainees' career perspectives after COVID-19: a qualitative study in China | to explore the Chinese GP trainees' career perspectives after COVID-19 utilizing a qualitative methodology |
| #67 | Factors associated with COVID-19 vaccine hesitancy in Senegal: a mixed study | to assess and identify factors associated with hesitancy toward the COVID-19 vaccine in Senegal |
| #68 | Factors associated with the acceptability of government measures to address COVID-19 in Senegal | to study the acceptability of government measures in Senegal concerning curfews, the prohibition of travel between regions, and the closure of markets and places of worship |
| #69 | Participating in a vaccine trial for COVID-19 in Senegal: trust and information | to understand the level and determinants of people's willingness to participate in a vaccine trial for COVID-19 in Senegal |
| #70 | Acceptability of government measures against COVID-19 pandemic in Senegal: A mixed methods study | to measure and understand the acceptability of these four governmental measures (the closure of places of worship, a curfew, a ban on movement between regions, and the closure of markets) as well as the level of public trust in the state to fight the pandemic |
